# Preparedness of health facilities for emergency neonatal care in the West Region of Cameroon: A hospital-based cross-sectional study

**DOI:** 10.64898/2026.11.23.26368724

**Authors:** Irene Ngassam Kamgno, Augustin Murhabazi Bashombwa, Merveille Claire Nana Djapou, Iyakachi Blama Abba, Hermann Aubin Ngounou Ntonwen, Collins Buh Nkum, Ange Vanel Wandji Nono, Ivan Walter Nguedia Dongmo, Cédrick Geordane Ateukeng, Jérome Ateudjieu

**Affiliations:** Department of Health Research, MA SANTE (Meilleur Accès aux soins de Santé), Yaoundé, Cameroon; Department of Public Health, Faculty of Medicine and Pharmaceutical Sciences, University of Dschang, Dschang, Cameroon; Faculty of Medicine, Université de Kaziba, Kaziba, Democratic Republic of Congo; Texila American University, Guyana, South America; Division of Health Operations Research, Ministry of Public Health, Yaounde, Cameroon

**Keywords:** emergency neonatal care, facility preparedness, neonatal mortality, West Region, Cameroon

## Abstract

**Background:** Limited access to quality emergency neonatal care (EmNC) contributes to high neonatal mortality in Cameroon, yet data on facility readiness in the West Region are scarce. This study assessed the preparedness of health facilities for EmNC in this region.

**Methods:** We conducted a cross-sectional study from March to May 2026 in five health districts of the West Region. All eligible facilities were included. Data were collected through a questionnaire for health professionals and an observation checklist adapted from the WHO/UNFPA SARA framework. Preparedness scores were classified as high (≥80%), moderate (60–79%) or low (<60%).

**Results:** We surveyed 185 health facilities (74.1% integrated health centers, 63.2% rural, 49.7% public) and 243 health professionals. The mean preparedness score was 51.1%; 11.4% of facilities had a high level of preparedness, 20.0% moderate and 68.6% low. At least one qualified staff member was present in 64.3% of facilities, and written neonatal protocols in 45.9%. Essential equipment was scarce: bag-valve-mask (13.0%), functional incubator (19.5%), newborn transport equipment (13.5%), phototherapy unit (10.8%) and defibrillator (7.6%), whereas vitamin K was widely available (95.7%). Mean scores were highest in private for-profit facilities (64.9%) compared with public (47.7%) and private non-profit facilities (45.1%), in district hospitals (66.2%) and CMAs (66.0%) compared with integrated health centers (45.9%), and in urban areas (60.9%) compared with rural (48.6%) and semi-urban areas (40.4%).

**Conclusions:** Most health facilities in the West Region are poorly prepared for newborn emergencies. Priorities include essential equipment with maintenance systems, qualified staff, written protocols and regular training, especially in integrated health centers and rural areas.

## Background

Limited access to quality Emergency Neonatal Care (EmNC) remains a major structural barrier that severely compromises newborn survival and exacerbates the socio-economic burden on families [1]. According to the World Health Organization (WHO), the neonatal period represents the most critical phase for child survival, accounting for 47% of all under-five deaths [2]. In 2022 alone, neonatal mortality caused an estimated 2.3 million deaths worldwide, with Sub-Saharan Africa bearing the heaviest burden, reporting a staggering mortality rate of 27 per 1,000 live births [3]. Cameroon reflects this regional crisis, with a national neonatal mortality rate of 28 per 1,000 live births [4]. However, this national average masks severe disparities, with reported mortality rates ranging from 18 per 1,000 live births in Buea to hospital-based case-fatality rates of 7.2% to 8.0% in urban referral centers like Yaoundé and Douala [5–7]. Evidence indicates that the primary drivers of these preventable deaths are neonatal asphyxia (66%), infections (37.8%), complications of prematurity (28%), and inadequate prenatal care visits by the mother [5–8].

In response to this critical situation, the WHO and the United Nations Children’s Fund (UNICEF) launched the “*Every Newborn: An Action Plan to End Preventable Deaths”*, advocating for core evidence-based interventions such as Kangaroo Mother Care, neonatal resuscitation, and healthcare provider empowerment [9]. In Africa, regional strategies use simulation training, standardized Emergency Obstetric and Neonatal Care (EmONC) kits categorized into two levels: Basic Emergency Obstetric and Neonatal Care (BEmONC) at health centers, and Comprehensive Emergency Obstetric and Neonatal Care (CEmONC) at hospitals, and task-shifting to improve emergency neonatal care [10]. Cameroon’s Ministry of Public Health and partners aligned with its 2021–2025 Strategic Plan, operationalize maternal and neonatal emergency care by deploying Health Voucher/ Universal Health Coverage (UHC) financial exemptions, regularly assessing facility needs, supplying EmONC kits, and continuously upskilling healthcare staff [11, 12]. However, in the West Region, only 30% to 40% of health facilities offer a basic EmNC package, which remains largely concentrated in urban referral hospitals [13]. This persistent mismatch between strategic policy and operational reality directly threatens newborn survival and underscores the urgent need to assess current EmNC capacity and implementation barriers in local healthcare settings [14].

Numerous studies across low and middle-income countries (LMICs) highlight critical systemic bottlenecks, including low overall service readiness (49%), severe equipment shortages (<25%), while demonstrating the clear benefits of standardized procedures and mothers who attended 4 or more antenatal care (ANC) visits had a 3.72-fold lower risk of infant mortality [15–19]. However, existing research in Cameroon has predominantly focused on clinical determinants and causes of neonatal mortality or remained restricted to single-center urban assessments [5–7]. Consequently, the actual level of readiness of health facilities to manage neonatal emergencies remains poorly documented, particularly across multi-district health systems [5–7]. To date, empirical data mapping the availability, equity, and functionality of essential human, material, and normative resources in the West Region remain lacking [13].

To address this evidence, this study provides the first comprehensive, multi-district evaluation in the West Region of Cameroon assessing the availability of essential human, material, and normative resources. Therefore, this study was proposed to assess the preparedness of health facilities for emergency neonatal care in the West region of Cameroon.

## Methods

### Study design

This was a cross-sectional study targeting health facilities providing EmNC in five health districts in the west region, Cameroon. Health districts were selected using purposive sampling, and all health facilities in the selected health districts were exhaustively selected. Data were collected using a face-to-face administered questionnaire to the health care professionals and a grid to assess the availability of recommended kit type per level of health facilities, equipment and consumables.

### Study setting and period

Data were collected from March to May 2026 in health facilities providing EmNC in Mifi, Dschang, Bangangté, Batcham, and Bandjoun health districts of the West region, Cameroon. The region comprises 20 health districts, serving an estimated total population of 2,384,678 inhabitants. This region is characterized by a high recorded birth rate of 46.8 births per 1,000 inhabitants per year, according to the 2018 Demographic and Health Survey (DHS) [14]. yet no data are currently available on the availability of emergency neonatal kits in these facilities, nor on the extent of neonatal deaths and complications. This region was chosen based on its high recorded birth rates, the limited evidence available in the literature on EmNC, and the lack of data on neonatal deaths and complications in the area.

### Study population

All health facilities providing EmNC that were formally registered in the Ministry of public health’ database in the selected districts were eligible. Health facilities that were permanently closed at the time of data collection and facilities that could not be reached or assessed for logistical or geographical reasons were not eligible. Those whose head consent to participate were included. These healthcare facilities included the maternity and pediatric departments whose department heads agreed to participate. Healthcare professionals from these selected departments were then included if they provided informed consent.

### Sampling process

The districts were stratified into urban, semi-urban, and rural areas, and health facilities were selected exhaustively. The urban district being considered as any district having more than three health areas in an urban setting; the semi-urban district with more than one health area in an urban setting and a rural district with one or less than one health area in an urban setting. Health facilities within these districts were further categorized as referral hospitals, District Hospitals, Medicalized Health Centers (CMA), and Integrated Health Centers (CSI), reflecting the different levels of the national health pyramid. Second, an exhaustive sampling approach was applied within each stratum using official health mapping lists. All eligible public, private, and confessional facilities offering EmNC services were systematically included to achieve the target sample size, ensuring balanced representation across geographic environments.

### Data collection tools and variable

Data were collected using a survey questionnaire and an observation checklist, both adapted from the WHO/UNFPA Service Availability and Readiness Assessment (SARA) framework and national EmNC guidelines [20]. A pretest was conducted in three health facilities to ensure the questions were clear and coherent for participants. The questionnaire collected information on the sociodemographic characteristics of the facilities, while the checklist gathered data on: (1) human resources and qualifications, (2) equipment, (3) essential medicines and medical supplies, and (4) normative documents and standard operating procedures.

### Data collection process

Eligible healthcare personnel were approached individually, informed about the study objectives and data collection procedures, and invited to participate. Written informed consent was obtained before any data were collected. Data were collected using a face to face administered questionnaire to assess information on the qualifications of the healthcare personnel.

After completing the assessment of healthcare personnel, the data collection team visited the departments providing emergency neonatal care to assess the availability of equipment, medical devices and supplies, consumables, essential medicines, and relevant clinical guidelines and normative documents. These items were assessed through direct on-site observation using the standardized observation checklist. All data collection activities were conducted by trained data collectors following standardized procedures

### Data management and data analysis

Data were collected via KoboToolbox and weekly supervisor checks ensured completeness, internal consistency, and inclusion adherence, with unresolvable entries treated as missing data, while non-respondents, refusals, non-consenting participants, and non-covered facilities were excluded. Descriptive statistics (frequencies, proportions with 95% CIs) summarized data in R software (version 4.3.0). EmNC preparedness scores were calculated based on the WHO/UNFPA SARA guidelines that have define signal functions [20]. Basic Emergency Obstetric and Newborn Care (BEmONC) covers 7 functions, including basic newborn resuscitation (airway suctioning, bag-mask ventilation, tactile stimulation, keeping the baby warm). Comprehensive Emergency Obstetric and Newborn Care (CEmONC) adds further functions such as full newborn resuscitation (suctioning, intubation, metabolic resuscitation), parenteral antibiotics and anticonvulsants for the newborn, and blood transfusion. These functions were operationalised using the resource- availability in our dataset, across two cumulative tiers: a BEmONC tier (11 items), applied to all HFs for the readiness score, and a CEmONC tier (20 items = BEmONC + 9 additional items), restricted to regional hospitals (n=2) for score calculation. For the descriptive tables below, however, item availability is reported across all 185 HFs for descriptive purposes, and stratified by facility type (public, private, confessional) and category (Regional Hospital, District Hospital, CMA, CSI) for disaggregated analysis. For the BEmONC package, a health facility has a high level of preparedness with 9 to 11 items (≥80%), a medium level with 7 to 8 items (60 - 79%), and low level with 6 items or fewer (<60%). For the CEmONC package, a health facility has high level of preparedness with 16 items to 20 items (≥80%), a medium level with 12 to 15 items (60 - 79%), and a low level with 11 items or fewer (<60%).

### Ethical Clearance

This study contributes to reducing neonatal mortality by identifying gaps in equipment and staffing. It strengthens health equity across districts and support training, NGOs, and researchers. Ethically it protects participants and ensures the fair dissemination of results. To minimize risks, free and informed consent was obtained from each participant, and data collection was anonymous and password-protected. Ethical clearance was granted by the Regional Ethics Committee for Human Health Research of the West Region (CRERSH-OU) under reference N° 442/25/02/2026/CE/CRERSH-OU/VP.

## Results

### Characteristics of participating health facilities

Table 1 and 2 present results on the characteristics of the health facilities and health professionals surveyed. Among the 185 health facilities, 137 (74.1%) was Integrated Health Centers (CSIs), 2 (1.1%) was Regional Hospitals. Public facilities represented 92 (49.7%) of the surveyed facilities, and 117 (63.2%) were located in rural areas. 192 (79.0%) of health professionals were female. 94 (38.7%) were State-registered nurses, followed by 49 (20.2%) midwives; and only 15 (6.2%) were physicians.

**Table 1:** Characteristics of the health facilities surveyed (n = 185)

| **Characteristic** | **Frequency (n)** | **Proportion (%)** | **95% CI** |
|---|---|---|---|
| **Facility type** |  |  |  |
| Public | 92 | 49.7 | 42.6 – 56.9 |
| Private for-profit | 44 | 23.8 | 18.2 – 30.4 |
| Private not-for-profit (faith-based/associative) | 49 | 26.5 | 20.7 – 33.3 |
| **Facility category** |  |  |  |
| Regional hospital | 2 | 1.1 | 0.3 – 3.9 |
| District hospital | 7 | 3.8 | 1.8 – 7.6 |
| Sub-divisional medical center (CMA) | 39 | 21.1 | 15.8 – 27.5 |
| Integrated health center (CSI) | 137 | 74.1 | 67.3 – 79.8 |
| **Setting** |  |  |  |
| Urban | 50 | 27.0 | 21.1 – 33.8 |
| Semi-urban | 18 | 9.7 | 6.2 – 14.9 |
| Rural | 117 | 63.2 | 56.1 – 69.9 |
| **Health district** |  |  |  |
| MIFI | 37 | 20.0 | 14.9 – 26.3 |
| DSCHANG | 46 | 24.9 | 19.2 – 31.6 |
| BANJOUN | 37 | 20.0 | 14.9 – 26.3 |
| BANGANTE | 37 | 20.0 | 14.9 – 26.3 |
| BATCHAM | 28 | 15.1 | 10.7 – 21.0 |

**Table 2:** Characteristics of health professionals surveyed (n = 243)

| **Characteristic** | **N** | **Proportion (%)** | **95% CI** |
|---|---|---|---|
| **Sex** |  |  |  |
| Female | 192 | 79.0 | 73.5 – 83.7 |
| Male | 51 | 21.0 | 16.3 – 26.5 |
| **Age group** |  |  |  |
| 18 to 21 years | 2 | 0.8 | 0.2 – 3.0 |
| 22 to 25 years | 34 | 14.0 | 10.2 – 18.9 |
| Over 25 years | 207 | 85.2 | 80.2 – 89.1 |
| **Education level** |  |  |  |
| Primary | 1 | 0.4 | 0.1 – 2.3 |
| Secondary | 47 | 19.3 | 14.9 – 24.8 |
| Tertiary/higher | 195 | 80.2 | 74.8 – 84.8 |
| **Professional qualification** |  |  |  |
| Physician | 15 | 6.2 | 3.8 – 9.9 |
| Senior nurse | 39 | 16.0 | 12.0 – 21.2 |
| State-registered nurse | 94 | 38.7 | 32.8 – 44.9 |
| Midwife | 49 | 20.2 | 15.6 – 25.7 |
| Lab technician | 13 | 5.3 | 3.2 – 8.9 |
| Other | 33 | 13.6 | 9.8 – 18.5 |

### BEmONC and CEmONC resources availability by facility category

Table 3 and 4 present the results on the BEmONC and additional CEmONC resources availability by facility category. 119 (64.3%) of health facilities has qualified and trained staff for neonatal emergency, 24 (13.0%) had bag-valve-mask, 36 (19.5%) had functional incubator, and 25 (13.5%) newborn transport equipment.

**Table 3:**
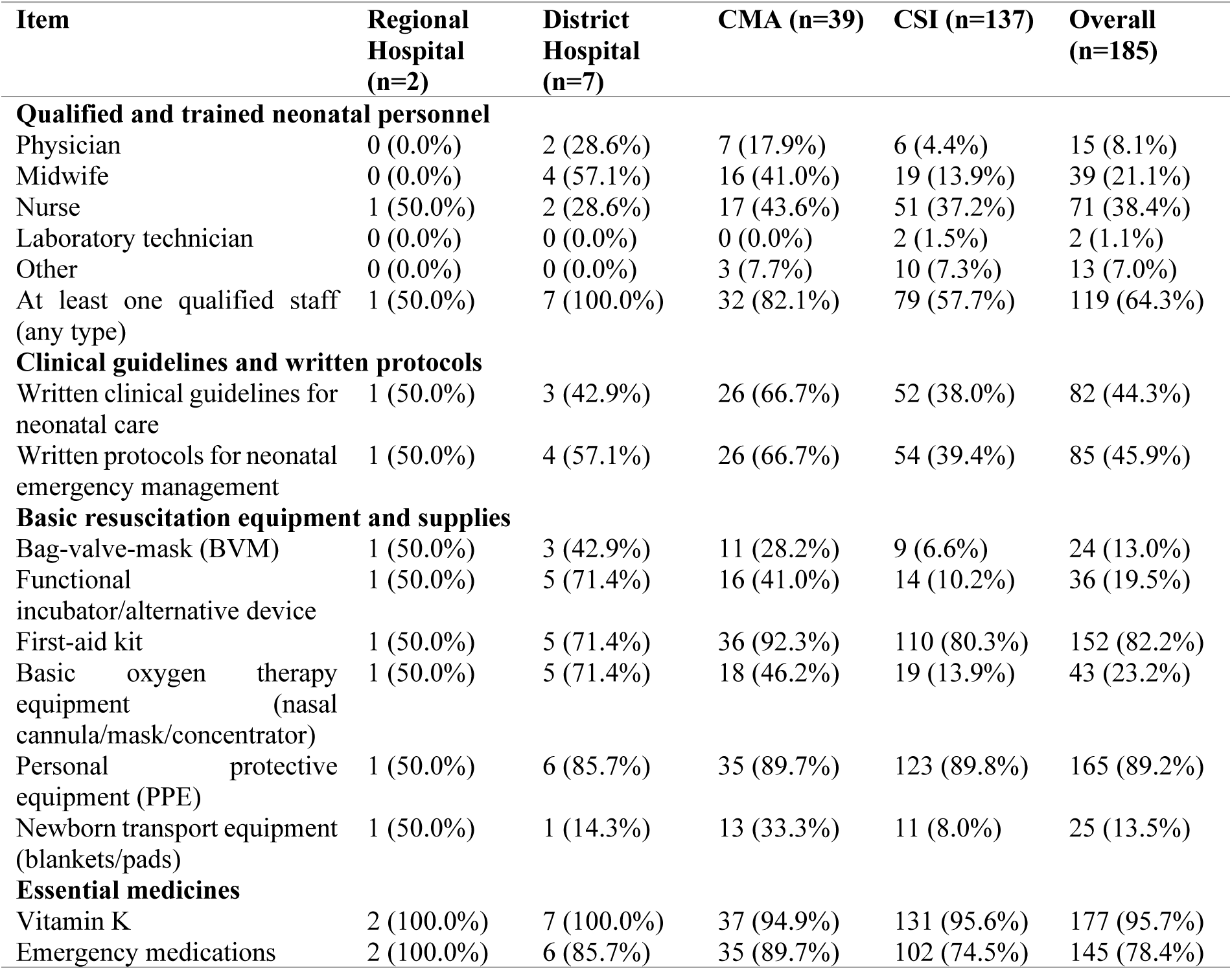
Availability of BEmONC resources by facility category

**Table 4:** Availability of additional CEmONC resources by facility category

| Item | Regional Hospital (n=2) | District Hospital (n=7) | CMA (n=39) | CSI (n=137) | Overall (n=185) |
|---|---|---|---|---|---|
| **Advanced monitoring** |  |  |  |  |  |
| Monitor (cardiac/vital signs) | 1 (50.0%) | 3 (42.9%) | 18 (46.2%) | 15 (10.9%) | 37 (20.0%) |
| Pulse oximeter | 1 (50.0%) | 5 (71.4%) | 20 (51.3%) | 24 (17.5%) | 50 (27.0%) |
| Defibrillator | 1 (50.0%) | 2 (28.6%) | 10 (25.6%) | 1 (0.7%) | 14 (7.6%) |
| **Advanced management** |  |  |  |  |  |
| Phototherapy unit | 1 (50.0%) | 3 (42.9%) | 10 (25.6%) | 6 (4.4%) | 20 (10.8%) |
| Emergency crash cart | 2 (100.0%) | 2 (28.6%) | 21 (53.8%) | 56 (40.9%) | 81 (43.8%) |
| Blood transfusion equipment/records | 2 (100.0%) | 7 (100.0%) | 28 (71.8%) | 31 (22.6%) | 68 (36.8%) |
| **Equipment sustainability** |  |  |  |  |  |
| Formal equipment maintenance system | 1 (50.0%) | 3 (42.9%) | 11 (28.2%) | 25 (18.2%) | 40 (21.6%) |
| Regularly receives training on resource utilization | 1 (50.0%) | 2 (28.6%) | 12 (30.8%) | 33 (24.1%) | 48 (25.9%) |
| **Advanced pharmacy** |  |  |  |  |  |
| BCG | 2 (100.0%) | 7 (100.0%) | 29 (74.4%) | 103 (75.2%) | 141 (76.2%) |
| Polio 0 | 2 (100.0%) | 7 (100.0%) | 25 (64.1%) | 90 (65.7%) | 124 (67.0%) |
| Hepatitis B vaccine | 2 (100.0%) | 7 (100.0%) | 28 (71.8%) | 62 (45.3%) | 99 (53.5%) |
| Prophylactic ARVs (newborn) | 2 (100.0%) | 6 (85.7%) | 24 (61.5%) | 52 (38.0%) | 84 (45.4%) |
| Caffeine | 2 (100.0%) | 3 (42.9%) | 11 (28.2%) | 9 (6.6%) | 25 (13.5%) |
| At least one item (composite indicator) | 2 (100.0%) | 7 (100.0%) | 36 (92.3%) | 112 (81.8%) | 157 (84.9%) |

Only 14 (7.6%) of health facilities had defibrillators and 20 (10.8%) phototherapy units, while emergency crash carts were available in 81 (43.8%) of facilities. Advanced pharmacy items, however, were available in 157 (84.9%) facilities.

### Preparedness of health facilities to provide EmNC in the West Region

Figure 1 present overall, the mean preparedness score was 51.1%: 21 of 185 facilities (11.4%) had a high level of preparedness, 37 (20%) a moderate level and 127 (68.6%) a low level.

**Figure 1:**
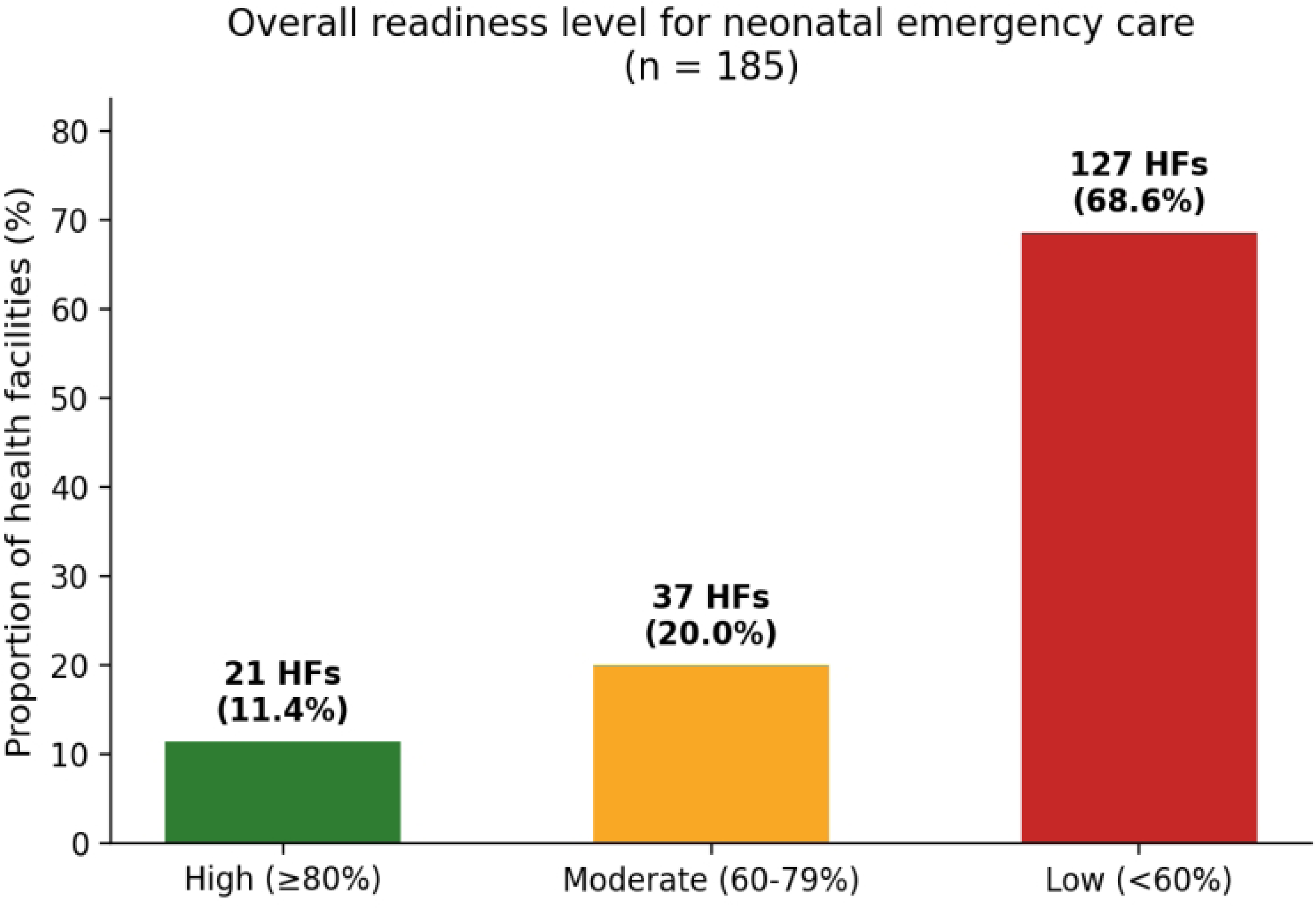
Overall preparedness level for neonatal emergency care

Table 5 and 6 present the preparedness scores by facility type and category. Private for-profit facilities had the highest mean score (64.9%) and the lowest proportion classified as low preparedness (40.9%), compare private non-profit (45.1%; 79.6% low) and public facilities (47.7%; 76.1% low). District Hospitals and CMAs had the highest mean scores (66.2% and 66.0%), while Integrated Health Centers (CSIs) had the lowest mean score (45.9%) and the largest proportion classified as low preparedness (79.6%).

**Table 5:** Preparedness score according to facility type

| **Facility Type** | Frequency (**n**) | **Mean Score (95% CI)** | **High (≥80%)** | **Moderate (60–79%)** | **Low (<60%)** |
|---|---|---|---|---|---|
| Private for-profit | 44 | 64.9% [58.8-70.9] | 12 (27.3%) | 14 (31.8%) | 18 (40.9%) |
| Private non-profit (confessional/faith-based/community) | 49 | 45.1% [39.1-51.0] | 4 (8.2%) | 6 (12.2%) | 39 (79.6%) |
| Public | 92 | 47.7% [44.1-51.3] | 5 (5.4%) | 17 (18.5%) | 70 (76.1%) |

**Table 6:** Preparedness score according to facility category (BEmONC/CEmONC level)

| **Health Facility Category** | Frequency **(n)** | **Mean Score (95% CI)** | **High (≥80%)** | **Moderate (60–79%)** | **Low (<60%)** |
|---|---|---|---|---|---|
| Regional Hospital | 2 | 62.5% [IC Inconclusive n=2] | 1 (50.0%) | 0 (0.0%) | 1 (50.0%) |
| District Hospital | 7 | 66.2% [55.4 - 77.0] | 2 (28.6%) | 2 (28.6%) | 3 (42.9%) |
| Sub-district Medical Center (CMA) | 39 | 66.0% [59.3 - 72.6] | 11 (28.2%) | 14 (35.9%) | 14 (35.9%) |
| Integrated Health Center (CSI) | 137 | 45.9% [42.9 - 49.0] | 7 (5.1%) | 21 (15.3%) | 109 (79.6%) |

Figure 2 present the preparedness score by setting. Urban facilities had the highest mean score (60.9%; 95% CI [54.5–67.2]), followed by rural (48.6%; [45.1–52.0]) and semi-urban facilities (40.4%; [30.3–50.5]). A high level of preparedness was reached by 24.0% of urban facilities, 8.5% of rural facilities and none of the semi-urban facilities, whereas 48.0% of urban, 76.1% of rural and 77.8% of semi-urban facilities had a low level.

**Figure 2:**
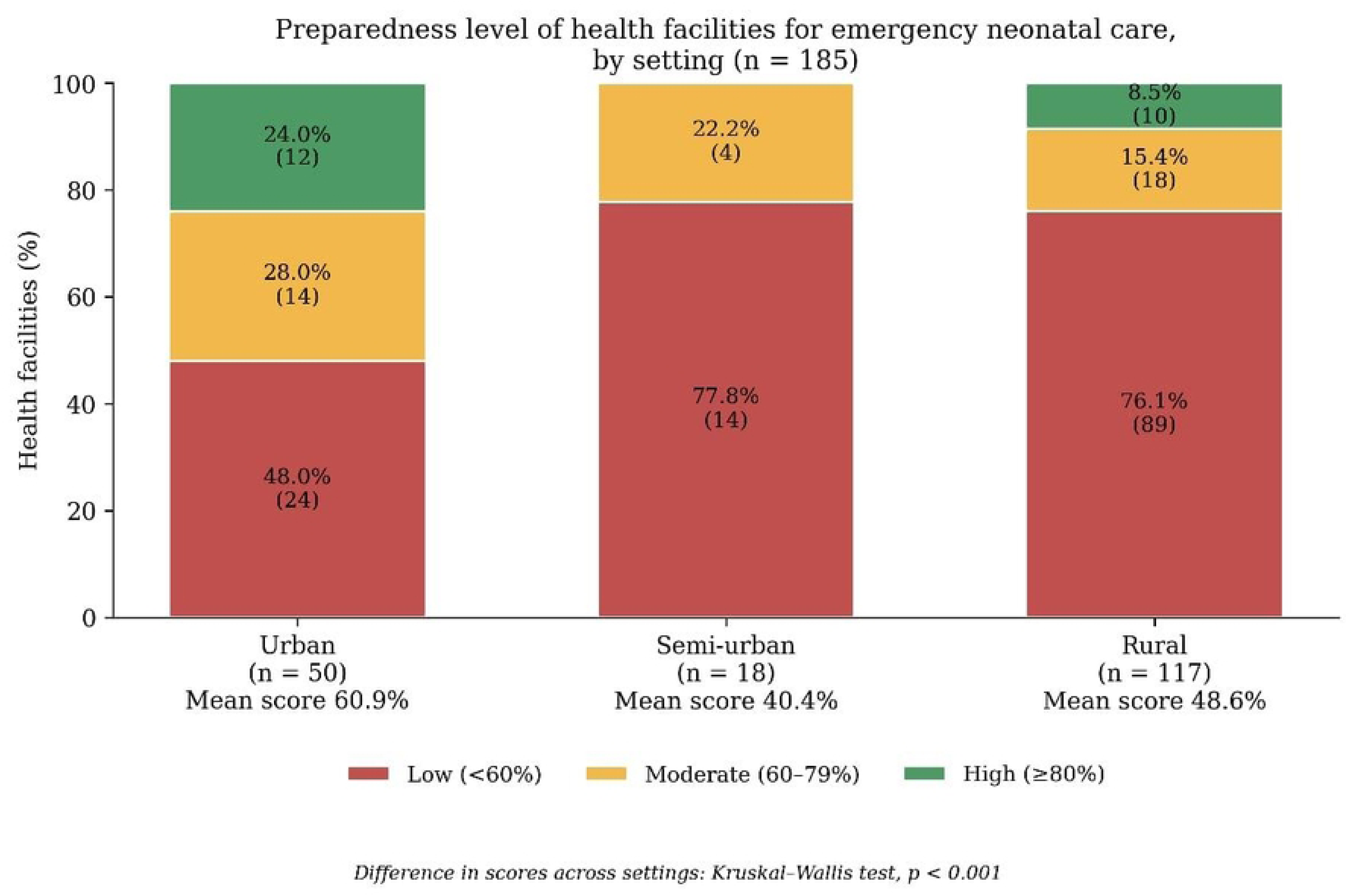
Preparedness level of health facilities for EmNC by setting.

## Discussion

This study assessed the preparedness of health facilities for emergency neonatal care in five health districts of the West Region of Cameroon. We surveyed 185 facilities. The mean preparedness score was 51.1%; 11.4% of facilities had a high level and 68.6% a low level. Qualified and trained staff were present in 64.3% of facilities, but written guidelines (44.3%) and protocols (45.9%) were often missing. Key equipment was rare: bag-valve-mask (13.0%), functional incubator (19.5%) and newborn transport equipment (13.5%), while vitamin K was widely available (95.7%). Advanced resources were also scarce, including defibrillators (7.6%), phototherapy units (10.8%) and formal equipment maintenance systems (21.6%). Private for-profit facilities scored highest (64.9%), compared with private non-profit (45.1%) and public facilities (47.7%). District Hospitals (66.2%) and Medicalized Health Centers (66.0%) scored higher than Integrated Health Centers (45.9%). Urban facilities scored highest (60.9%), followed by rural (48.6%) and semi- urban facilities (40.4%).

Overall, preparedness for emergency neonatal care (EmNC) was suboptimal across the participating health facilities, with a mean preparedness score of 51.1%. This finding aligns broadly with studies from low- and middle-income countries, which report overall readiness levels of approximately 49% and availability of essential equipment below 25% [15–19]. The mean preparedness score observed in our study, along with the limited availability of key resources such as bag-valve-mask devices, incubators, and oxygen equipment (ranging from 13.0% to 23.2%), falls within a similar range. These findings highlight significant gaps in human resources, equipment, and other essential inputs required for the timely management of neonatal emergencies. Such deficiencies are inconsistent with WHO and UNFPA recommendations, which state that facilities providing EmNC should have appropriately qualified personnel and the essential equipment necessary for basic emergency obstetric and neonatal care. Addressing these shortcomings is therefore critical to improving the safety and quality of neonatal care in the region. These findings may also help explain the high burden of severe neonatal conditions reported in some health facilities in Cameroon, where neonatal asphyxia, infections, and complications of prematurity are among the leading causes of neonatal death, with reported case-fatality rates ranging from 7.2% to 8.0% [5–8]. The limited availability of resources required to recognize and manage these conditions in many healthcare facilities included in this study suggests that some newborns may be referred without adequate initial stabilization. Delayed referral or inadequate stabilization before transfer could potentially contribute to poor outcomes at the referral level, although this pathway was not directly assessed in our study. Strengthening the readiness of first- contact facilities, including ensuring the availability of appropriately trained personnel, essential equipment, and clinical guidelines, may therefore represent a crucial component of efforts to improve the continuum of neonatal care. Future studies should assess whether facility-level EmNC preparedness is associated with referral patterns, quality of stabilization before transfer, and neonatal survival.

Only 64.3% of facilities had qualified and trained staff, and physicians (4.4%) and midwives (13.9%) were scarce in Integrated Health Centers. A survey of 84 facilities in Malawi, which also assessed the presence of staff trained in emergency obstetric and newborn care, found that 75% of health centers had no clinician trained and 39% had no trained midwife [21]. Hospitals were also affected, with 62% having no trained clinician and 15% no trained midwife [21]. The situation in our facilities appears less severe, as about one in three had no qualified and trained staff. This difference may be explained by our broader definition, which counted staff of any cadre, whereas the Malawian survey considered only clinicians and midwives trained in basic emergency obstetric and newborn care. The surveys also took place in different countries and years. Despite this difference, both studies show that, in many primary-level facilities, no staff member has the training needed to manage a newborn emergency. This is worrying because the Ethiopian national survey found that providers’ knowledge of neonatal resuscitation was higher among trained providers and in facilities with a guideline [15]. In addition, in our study fewer than half of the facilities had written guidelines or protocols, only 25.9% received regular training on the use of their resources, and only 21.6% had a system to maintain equipment. Studies in other low-resource settings show that standard procedures combined with staff training improve the quality of care [18]. The situation is also far from Cameroon’s strategic plan, which aims to supply EmONC kits and to train health staff continuously [11, 12]. We recommend that the Regional Delegation of Public Health and district managers prioritize the deployment of qualified staff to Integrated Health Centers, the distribution of neonatal protocols, and regular in-service training combined with equipment maintenance.

This study has limitations; first the five districts were purposively selected to represent the rural, urban and semi-urban settings of the West Region, the results can therefore be considered representative of the West Region, but they cannot be generalized to the whole of Cameroon, where health system organization and resources may differ. Second, the score measures whether resources were available at one point in time. It does not measure how well they work over time; supply interruptions, quality of care or staff performance, and the cross-sectional design does not allow conclusions on neonatal outcomes. Third, staff qualifications were self-reported, which may cause reporting bias. Fourth, the score gives the same weight to all items and uses fixed cut-offs, so it cannot be compared directly with other readiness indices. Finally, the results for Regional Hospitals (n = 2) and semi-urban facilities (n = 18) are imprecise, and differences by facility type, category and setting were not adjusted for each other.

## Conclusion

This first multi-district assessment shows that most health facilities in the West Region of Cameroon are poorly prepared to manage newborn emergencies. The mean preparedness score was 51.1%, and only 11.4% facilities reached a high level. Lifesaving basics were often missing, only 13% of health facilities had bag-valve-masks, 19.5% had a functional incubator and 45.9% had written protocols. The gaps were largest in Integrated Health Centers, public and faith-based facilities, and rural areas. These findings highlight the need for health system and health facility managers to i) equip every facility providing newborn care with a minimum set of essential equipment, including bag-valve-masks, incubators and newborn transport equipment, and set up a system to maintain it; ii) ensure the presence of qualified staff, written protocols and regular in- service training, especially in Integrated Health Centers; and; iii) give priority to rural, public and faith-based facilities, where the gaps were largest, and monitor progress regularly.

## Data Availability

All relevant data are within the manuscript and its Supporting Information files. Additional anonymized datasets used during the study are available from the corresponding author upon reasonable request.

